# Perspectives of Key Stakeholders on Integrating Wearable Sensor Technology into Rehabilitation Care: A Mixed-Methods Analysis

**DOI:** 10.1101/2024.11.25.24317911

**Authors:** Allison E. Miller, Carey L. Holleran, Marghuretta D. Bland, Ellen E. Fitzsimmons-Craft, Caitlin A. Newman, Thomas M. Maddox, Catherine E. Lang

## Abstract

**Introduction:** Rehabilitation is facing a critical practice gap: Patients seek out rehabilitation services to improve their activity in daily life, yet recent work demonstrates that rehabilitation may be having a limited impact on improving this outcome due to lack of objective data on patients’ activity in daily life. Remote monitoring using wearable sensor technology is a promising solution to this address this gap. The purpose of this study was to understand patient and clinician awareness of the practice gap and preferences for integrating wearable sensor technology into rehabilitation care.

**Methods:** This study used a mixed-methods approach consisting of surveys and 1:1 interviews with clinicians (physical and occupational therapists or assistants) employed at an outpatient rehabilitation clinic within an academic medical center and patients seeking care at this clinic. Data were analyzed using descriptive statistics and thematic analysis.

**Results:** Data saturation was reached from recruiting nineteen clinicians and ten patients. Both clinicians and patients recognized the importance of measuring activity outside the clinic and viewed wearable sensor technology as an objective measurement tool. Most clinicians (63%) preferred continuous (vs. intermittent) monitoring within a care episode and most patients (60%) were willing to sync their sensor data as often as instructed by their provider. To maximize integration into clinical workflows, clinicians voiced a preference for availability of sensor data in the electronic health record.

**Conclusions:** Clinicians and patients value the use of wearable sensor technology to improve measurement of activity outside the clinic environment and expressed preferences for how this technology could best be integrated into routine rehabilitation care.

## INTRODUCTION

Rehabilitation clinicians (e.g., physical and occupational therapists) are key providers for improving movement and overall function in individuals with disabilities. One of the most common reasons patients seek out rehabilitation services is to improve their activity in daily life.(1, 2) The brief, episodic nature of clinic visits, however, makes it difficult to obtain a comprehensive and accurate picture of patients’ activity in daily life. In-clinic measures of a person’s *capacity* for activity (i.e., what a person *can do* measured by standardized assessments) have historically been used as a surrogate measure of what a person *actually does* (activity *performance*) in daily life.(3) This assumption posits that if a person’s capacity for activity is improved in the clinic that they will have resultant improvements in their activity performance in daily life.(4) Results from several seminal studies, however, have disproven this assumption and reinforced that activity capacity and activity performance are different constructs.(5–8)

A trial of individuals with chronic stroke randomized participants into one of three interventions groups to determine which intervention would yield the greatest change in activity performance (measured by steps/day using an activity tracker): a high-intensity walking intervention aimed at improving activity *capacity*, a step activity monitoring behavioral intervention aimed at improving activity *performance*, or a group that received *both* interventions.(8) Results showed that different interventions are required to improve an individual’s activity capacity versus activity performance in daily life and that improvements in activity capacity were not necessarily accompanied by improvements in activity performance in daily life (see Table 3 in Thompson et al., 2024).(8) A longitudinal cohort study of individuals with stroke and Parkinsons disease enrolled in outpatient rehabilitation care in five sites across the United States demonstrated that this discrepancy is not unique to the research realm but exists in actual clinic practice in which the majority (59%) of individuals improved in their activity capacity but not in their activity performance in daily life.(5) A secondary analysis of data from this study provide a possible explanation for these findings in which only 21% of clinician-documented goals were aimed at improving activity performance in daily life.(9) These data highlight a critical practice gap between the goals of patients (improving activity *performance* in daily life) and what is typically measured and intervened on in rehabilitation practice (activity *capacity*) and further suggest that rehabilitation may not yet be optimized for improving outcomes most salient to patients.(1, 2)

Addressing this practice gap, like many other practice gaps in rehabilitation (10–12) and other realms of medicine (13, 14), will require a targeted, multidisciplinary approach among researchers, clinicians, implementation scientists, and clinical leadership. These implementation efforts, however, will be ineffective without an understanding of why the practice gap exists.(15) One potential explanation for this practice gap is whether or not clinicians are aware of the gap between activity capacity and activity performance. If clinicians are not aware of the practice gap, then efforts to address this gap in clinical practice will fall short. A second potential explanation is that the tools needed to directly measure activity performance in daily life, wearable sensors, are not available or scantly used in most rehabilitation clinics.(4, 16) Rehabilitation clinicians have historically relied on self-report measures of activity performance in daily life, but previous work has shown that a person’s self-report of their activity performance in daily life is neither consistent nor accurate compared to direct (sensor-based) measures.(17–19) A third potential reason for this practice gap is the lack of technology integration into clinical workflows, a common reason why many digital health interventions are not adopted in healthcare settings.(20–22) To explore these potential reasons, stakeholder engagement is needed to: (1) understand awareness of the problem, (2) identify key barriers and facilitators that should be considered in implementation protocols, and (3) design workflows that seamlessly integrate the technology into rehabilitation practice and pose minimal burden on stakeholders.(15, 23)

While previous studies have investigated clinician and patient perceptions on wearable sensors (24, 25) and digital health technology more generally (26–28), no studies have investigated clinician and patient perspectives on using wearables sensors to monitor activity performance in daily life within a rehabilitation care episode. To explore these perspectives and facilitate efforts that seek to integrate wearable sensor technology into rehabilitation practice, we employed a mixed methods approach consisting of surveys and one-on-one interviews with patients and clinicians to answer four research questions (RQ):

RQ1. How do clinicians perceive the constructs of activity capacity and activity performance and their relationship in outpatient rehabilitation?

RQ2. How do clinicians and patients perceive the value of activity performance monitoring in outpatient rehabilitation care?

RQ3. What approaches, if any, are currently being used to measure activity performance in daily life in outpatient rehabilitation practice, and what are important considerations regarding these approaches?

RQ4. What are the data collection and workflow preferences of clinicians and patients for integrating wearable sensor technology into rehabilitation care?

Data from each of these questions were then used to develop a conceptual model that described the various approaches to activity performance monitoring within outpatient rehabilitation practice and their respective barriers and facilitators.(29)

## METHODS

### Participants

Licensed physical and occupational therapist clinicians employed within Washington University’s outpatient rehabilitation system were recruited using targeted email blasts and flyers. Targeted emails helped ensure the clinician sample was representative of both physical and occupational therapists who were treating a variety of movement problems resultant from both neurological and musculoskeletal diagnoses, and that the sample included clinicians with a range of experience using activity performance monitoring in their practice. Patients were recruited via clinician referral and included if they met the following criteria: (1) currently being seen in outpatient rehabilitation at Washington University for upper and/or lower limb problems, (2) access to a mobile phone, and (3) no significant cognitive or communication deficits that would limit their ability to participate in an interview with a research team member. Access to a mobile phone was required, as some of the interview questions asked about preferences for syncing data via a mobile app. Washington University Human Research Protection Office approved the study, and all participants signed informed consent prior to engaging in study activities.

### Procedures

All participants completed two surveys, a demographic survey and a survey evaluating their experiences with wearable sensor technology, and a one-on-one interview. Participants were provided the option to complete the surveys electronically using REDCap (Research Electronic Data Capture) (30, 31) or in-person just prior to the start of the interview. Interviews were semi-structured, approximately 45 minutes in length, and completed in person (in an outpatient rehabilitation clinic) or via Zoom.

Patient-specific and clinician-specific semi-structured interview guides were created, reviewed, and revised by study team members, which included rehabilitation clinicians, researchers, and educators, until consensus was achieved. Interview guides included questions exploring global reflections on concepts related to wearable sensor technology and our local efforts to integrate wearable sensors into rehabilitation practice. Table 1 summarizes the quantitative and qualitative question response data that was used to answer each research question. Prior to the first interview, an in-service was provided to clinicians to discuss the project and answer any questions. All interviews were conducted by two of the authors (AEM or CLH) and audio recorded and transcribed using Zoom. To ensure accuracy in the transcribed data, a research team member conducted a thorough review of all interview transcripts against the original audio recordings and made necessary corrections prior to analysis.

**Table 1.**
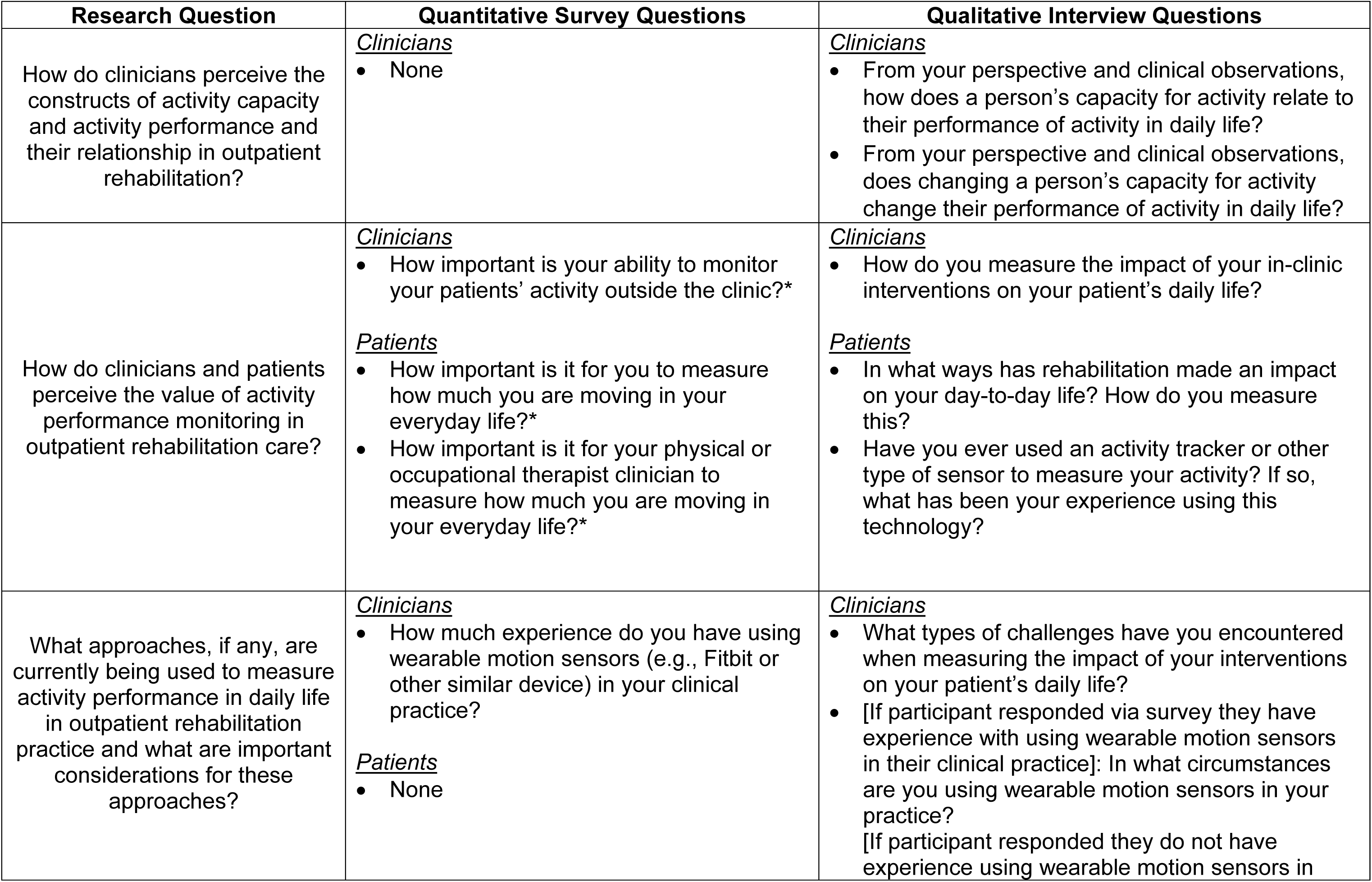
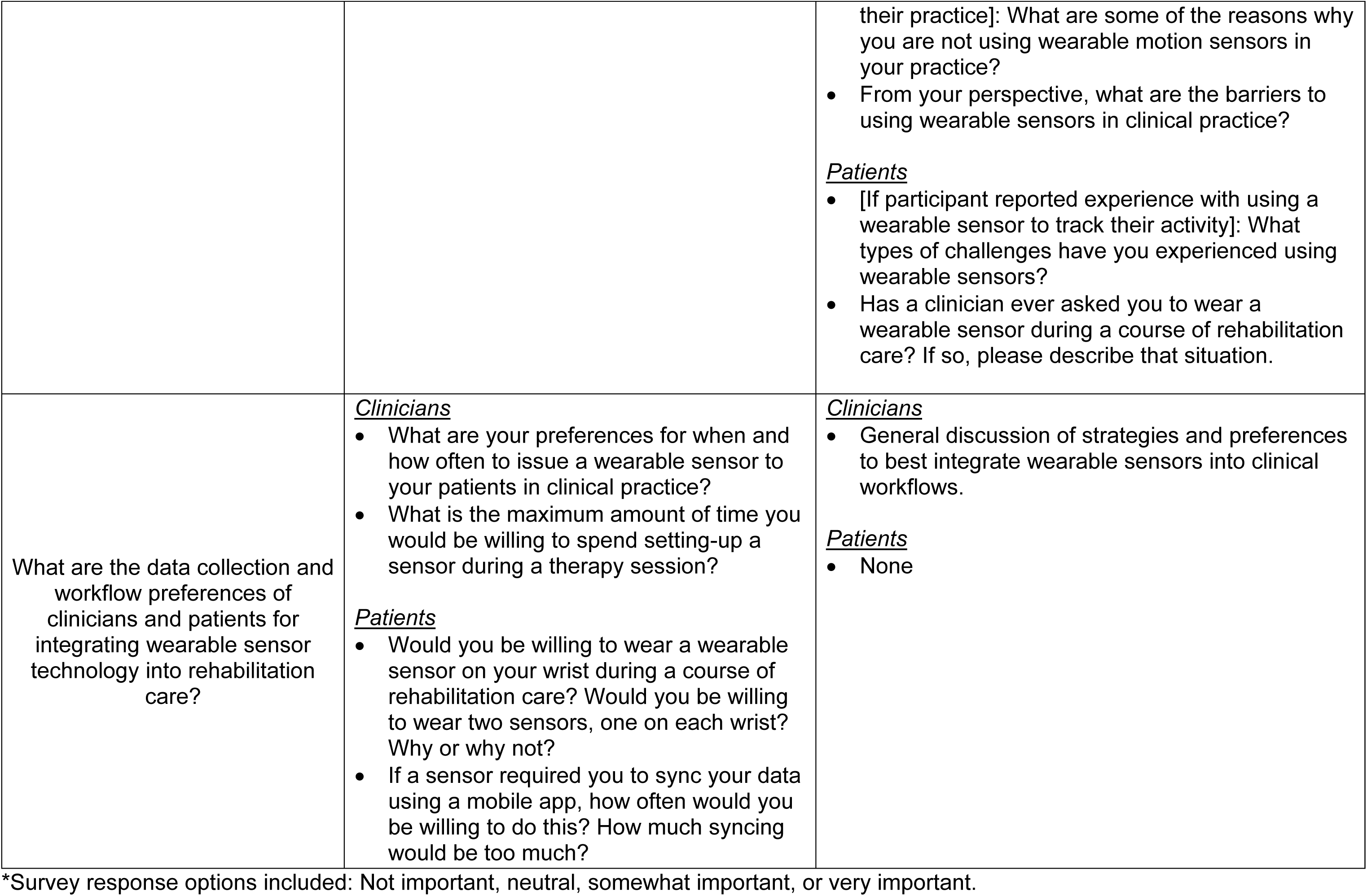
Quantitative and Qualitative Data Used to Address Each Research Question.

### Data Analysis

Survey response data were analyzed using descriptive statistics in R (R Core Team 2021, version 4.2.1).(32) Interview data were analyzed through a deductive and inductive thematic approach depending on the research question.(33, 34) Each interview was independently coded by two researchers, AEM and CLH, using the qualitative research software, HyperRESEARCH. Each clinician and patient’s narrative transcript served as a “case” for analysis. For RQ1, AEM and CLH separately coded clinicians’ responses to determine if their description of the constructs of activity capacity and activity performance matched published definitions (3) in order to quantify whether the clinician did or did not have an understanding of these constructs as they relate to their clinical practice. A third researcher, MDB, was used when there was a discrepancy between the two primary coders. For RQ 2-4, the initial coding process involved inductively identifying codes based on the clinician’s and patient’s own words as they related to each research question.(33, 34) After independently coding 3-5 interviews, AEM and CLH held meetings to discuss and group similar codes, refining and creating a revised code book to guide further coding for each research question.

For example, in regard to RQ3 exploring approaches to monitoring activity performance in daily life, “cost/access” and “insurance/reimbursement concerns” were highlighted as two separate considerations brought up by two different participants. These two codes were combined to generate the theme “Cost/access considerations for patients to purchase a wearable sensor”. This iterative process continued as AEM and CLH independently coded additional sets of 3-5 transcripts, followed by meetings to discuss, revise, and consolidate codes into themes organized by each research question. This iterative coding, revision, and theme generation process continued until all cases were coded and new codes related to each research question ceased to emerge. Clinicians and patients were recruited until data saturation was reached and no new codes were gleaned from additional interviews. After all interviews had been analyzed, AEM and CLH, met to ensure a consolidated list of themes by research question had been developed. The final list of themes as organized by research question and overall conceptualization of the data was presented to the research team for feedback. A conceptual model was then generated to display the process, pitfalls, and opportunities of differing methods of measuring activity performance in daily life within outpatient rehabilitation practice.

## RESULTS

Recruitment occurred from September 2023 through March 2024. Twenty-two occupational and physical therapist clinicians were contacted, 21 were screened for eligibility (1 did not respond to the request to complete eligibility questions), and 19 were consented and completed all study activities (1 did not complete the informed consent document and 1 declined to participate). Fifteen patients were referred from clinicians and 10 were screened, consented, and completed all study activities (5 did not respond after being contacted by the research team).

Clinician experience using wearable motion sensors in their practice ranged from 0 to 6+ years. Six patients were currently using a wrist-worn sensor or mobile app to monitor their activity, two had previous experience using technology to monitor their activity but were not currently doing so, and two did not have any experience. Tables 2 and 3 displays descriptive statistics for the clinician and patient cohorts, respectively.

**Table 2.**
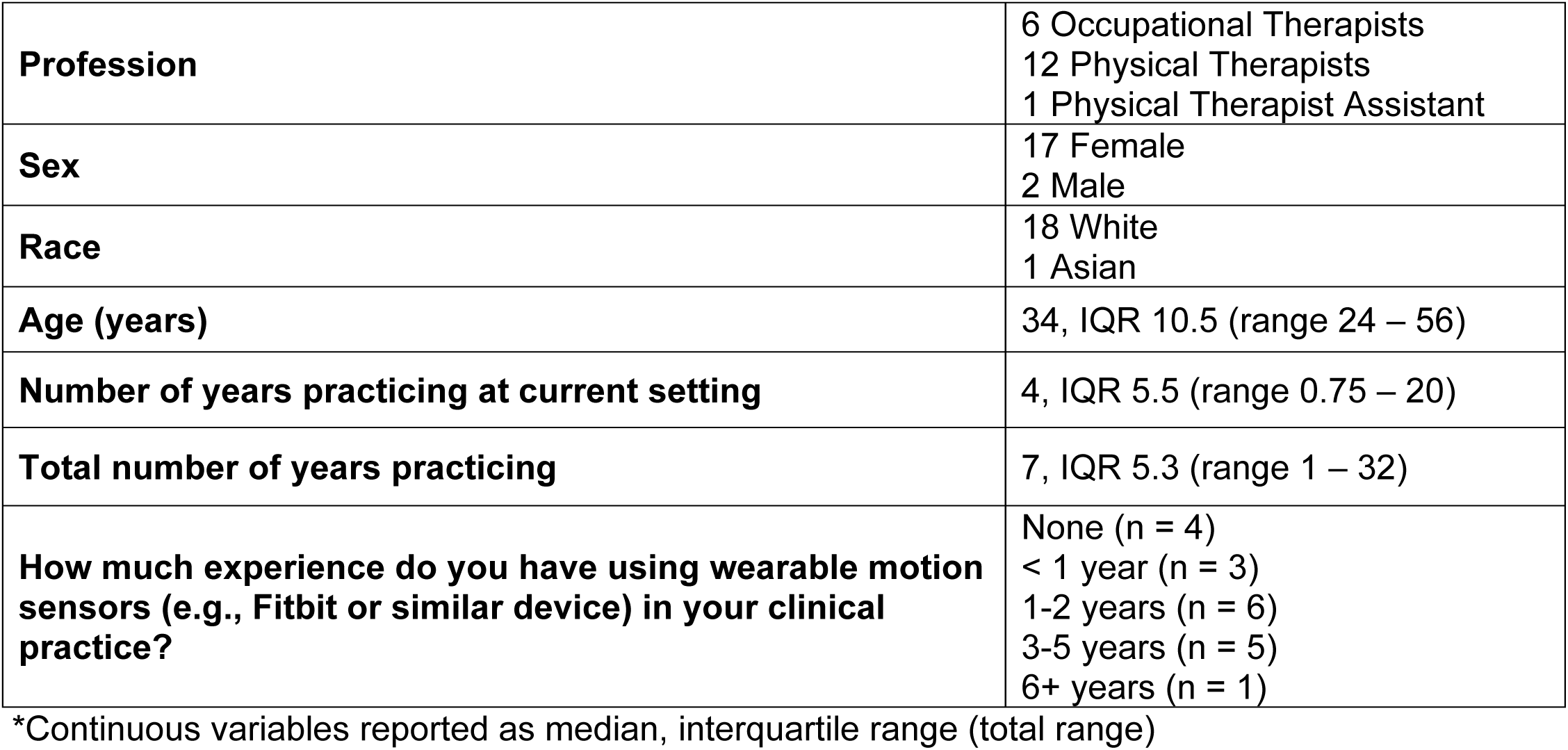
Demographic Characteristics of the Clinician Cohort (n = 19)*

**Table 3.**
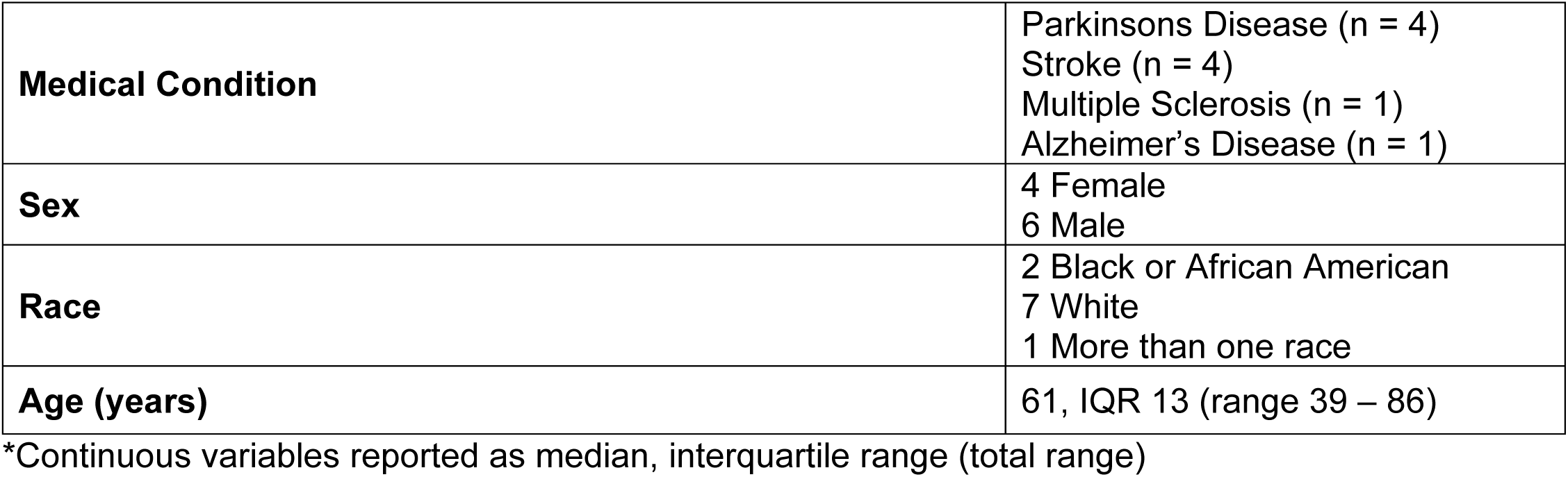
Demographic and Clinical Characteristics of Patient Cohort (n = 10)*

### Research Question 1. How do clinicians perceive the constructs of activity capacity and activity performance and their relationship in outpatient rehabilitation?

Adjudication by a third reviewer (MDB) was required on three cases to determine whether clinicians understood the distinction between activity capacity and activity performance. This resulted in sixteen out of nineteen clinicians (84.2%) who accurately described the difference between activity capacity and activity performance during the interview. The remaining three clinicians (15.8%) did not accurately describe the distinction between these two constructs. In the below excerpt, a clinician demonstrates understanding of activity capacity, by example of the 6-minute walk test, and activity performance, by example of a patient increasing their day-to-day activity:

> “I’ve also had people that, you know, they make a lot of progress on test measures like [the 6-minute walk test] as they’re working with me and they are also getting more active in their day-to-day, too.” – Clinician 4

When sharing their perceptions of the relationship between activity capacity and activity performance in daily life, several clinicians voiced that in-clinic assessments of activity capacity provide only a “snapshot” of the patient and are not necessarily informative of the patient’s activity performance in daily life.

> “I think based on my clinical observations, we are only really getting a snippet of things when we’re taking our [in-clinic] measures…Yes, we may see some improvements on [in-clinic measures], but I don’t know that is always getting the full picture of [the patient] outside the clinic. That is where there’s kind of a gap of like how do we monitor something like that. Sometimes subjectively we can get that as far as like they feel like they’ve done more, or they walk around longer in a store… but to actually have something quantifiable, there is I think a little bit of a gap there.” – Clinician 3

> “Some patients, you know, their increase in activity level is definitely reflected in their 6-minute walk test and vice-versa, and other times it’s not… Sometimes they might do really well in the clinic, because they’re motivated, they’re kind of tested on this measure and they know, this is gonna get sent to my doctor or my family member is gonna to read this note, and so…they do really well, but then I know at home they’re not very active from their report or their caregiver’s report.” – Clinician 8

Although some clinicians could not accurately distinguish the constructs of activity capacity and activity performance, all clinicians acknowledged the limitations of brief, in-clinic assessments for understanding a patient’s activity performance in daily life.

### Research Question 2: How do clinicians and patients perceive the value of activity performance monitoring in outpatient rehabilitation care?

When surveyed, twelve clinicians responded that it is “very important” to be able to monitor their patients’ activity outside of clinic sessions, with six clinicians reporting this is “somewhat important”, and one clinician who was neutral on this issue (Figure 1, left panel). Patient perceptions were generally similar, with the majority (n = 6) of patients responding it is “very important” to measure how much they are moving in daily life and for their physical and/or occupational therapist to be able to measure this (n = 8; Figure 1, middle and right panels). Patients expanded on these concepts during interviews, in which several patients who were currently using a wearable sensor reported that having the device helps them stay accountable with their activity.

> “And if I’m getting lazy or tired and I put [activity] off, then I’ll look at [the sensor] and say ‘no, I got to get going’, and I will get going and do it.” – Patient 28

**Figure 1.**
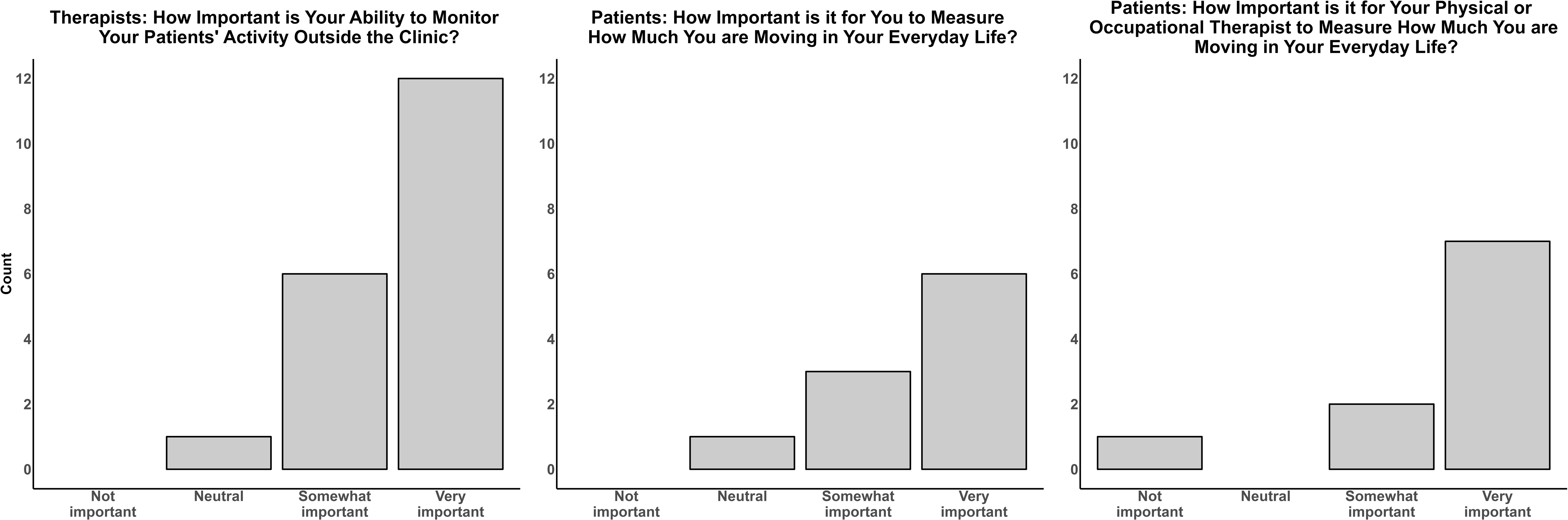
Perceived importance of Activity Performance Monitoring by Clinicians and Patients. The left bar graph displays clinician ratings on the importance of measuring their patients’ activity outside the clinic. The middle bar graph displays patient ratings on the importance of measuring their own activity in daily life. The right bar graph displays patients’ perceived importance of their therapist clinician to be able to measure their activity in daily life.

In addition to personal accountability, some patients reported that wearing a wearable sensor during rehabilitation care could also help them stay accountable to their physical and/or occupational therapist.

> “I think it would be helpful, because if I didn’t do it, I know [my therapist] is gonna be watching.” – Patient 25

Overall, the majority of clinicians and patients valued activity performance monitoring in outpatient rehabilitation care.

### Research Question 3. What approaches, if any, are currently being used to measure activity performance in daily life in outpatient rehabilitation practice and what are important considerations regarding these approaches?

Clinicians described two main approaches to monitoring activity performance in outpatient rehabilitation care: (1) the use of self-report measures or activity logs, and (2) consumer-grade wearable sensors owned or acquired by the patient. Almost all clinicians mentioned using self-report measures or activity logs to measure their patients’ activity performance in daily life. Clinicians expressed several limitations with using self-report measures or activity logs including high patient burden and questionable reliability of information. Consequently, clinicians voiced that this approach often results in data loss and inaccurate or unusable data.

> “If [a patient] is doing a walking program and it’s more just self-report, okay, well, is that accurate? Are [they] just telling me what I want to hear?” – Clinician 1

Seventy-nine percent (15/19) of clinicians reported having some experience with using consumer-grade wearable sensors with patients who already owned or were willing to purchase a device. Clinicians expressed that this approach was often more desirable than measuring activity performance using self-report measures as it increased reliability of the information by having quantifiable data from a wearable sensor.

> “I think [using a wearable sensor] helps with the confidence [of my clinical decisions] at least because it gives me more objective information of what they’re actually truly doing outside of the clinic…So that changes the education piece at least and having that data really helps because, like I said, sometimes it’s not even that they’re not trying to be honest or forthcoming with information, they just don’t realize how little they’re [the patient] is actually moving.” – Clinician 4

Despite finding that most clinicians perceived activity performance monitoring as “somewhat” or “very important” and reported experience using consumer-grade sensors with their patients, only 40% of patients reported using a consumer-grade sensor in collaboration with their therapist clinician during outpatient rehabilitation care. This finding may be related to important themes for consideration raised by clinicians and patients for integrating wearable sensors into rehabilitation care (Tables 4 and 5, respectively). These considerations could be viewed as a barrier or facilitator, depending on the context. For example, the cost for patients to purchase a wearable sensor was reported as a barrier if the patient did not have the financial means. It was reported as a facilitator, however, when the patient was able to purchase a device and because of the increasing number of consumer devices available at lower price points.(35) Considerations voiced by both clinicians and patients included cost, comfort with technology, presence of patient impairments that may impact adherence to remote performance monitoring, and trust (or lack-there-of) in the accuracy of the sensor.

**Table 4.**
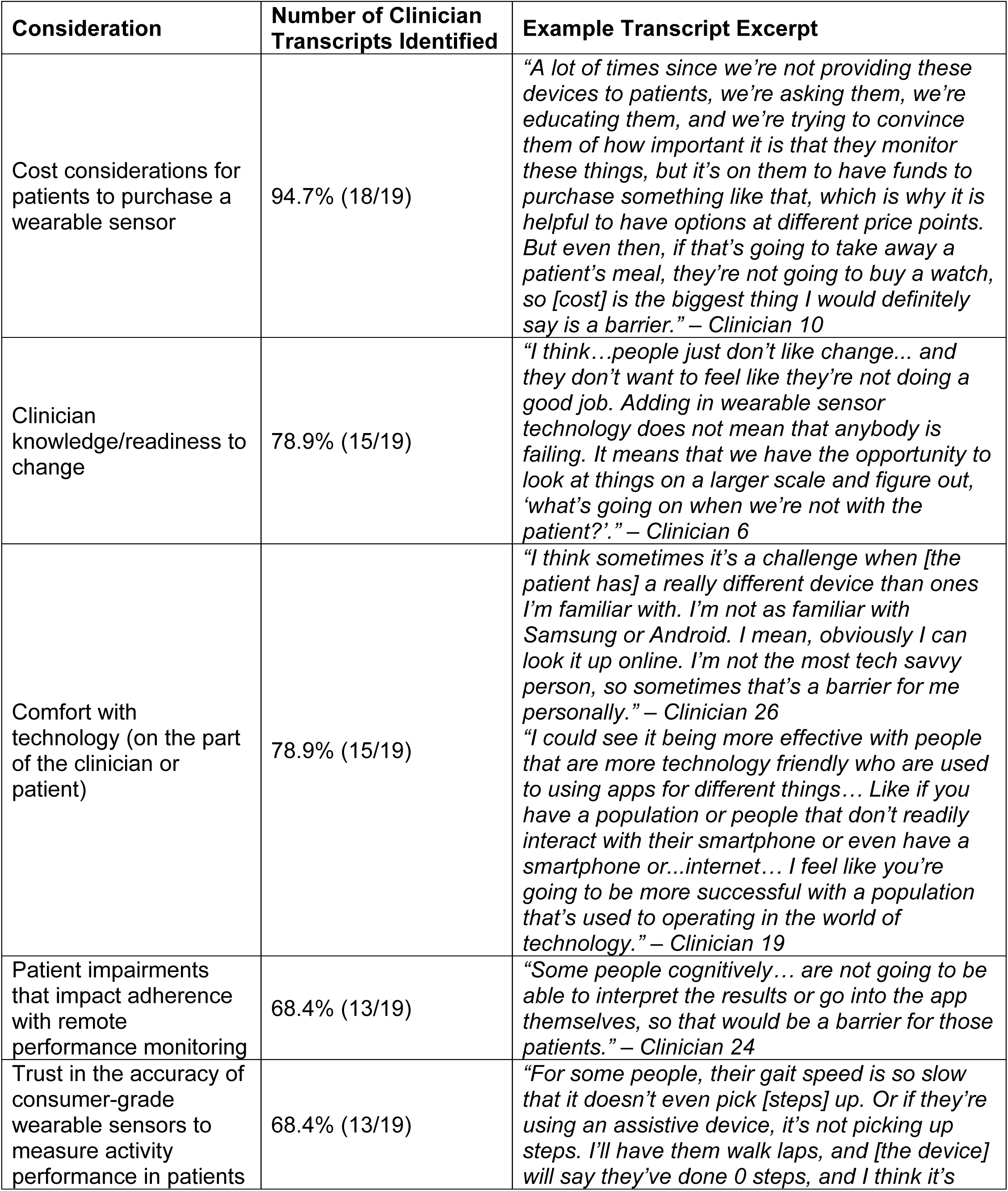
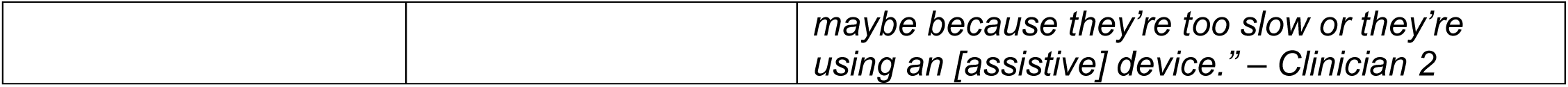
Top Five Clinician-Reported Considerations to Integrating Wearable Sensor Technology into Rehabilitation Care.

**Table 5.**
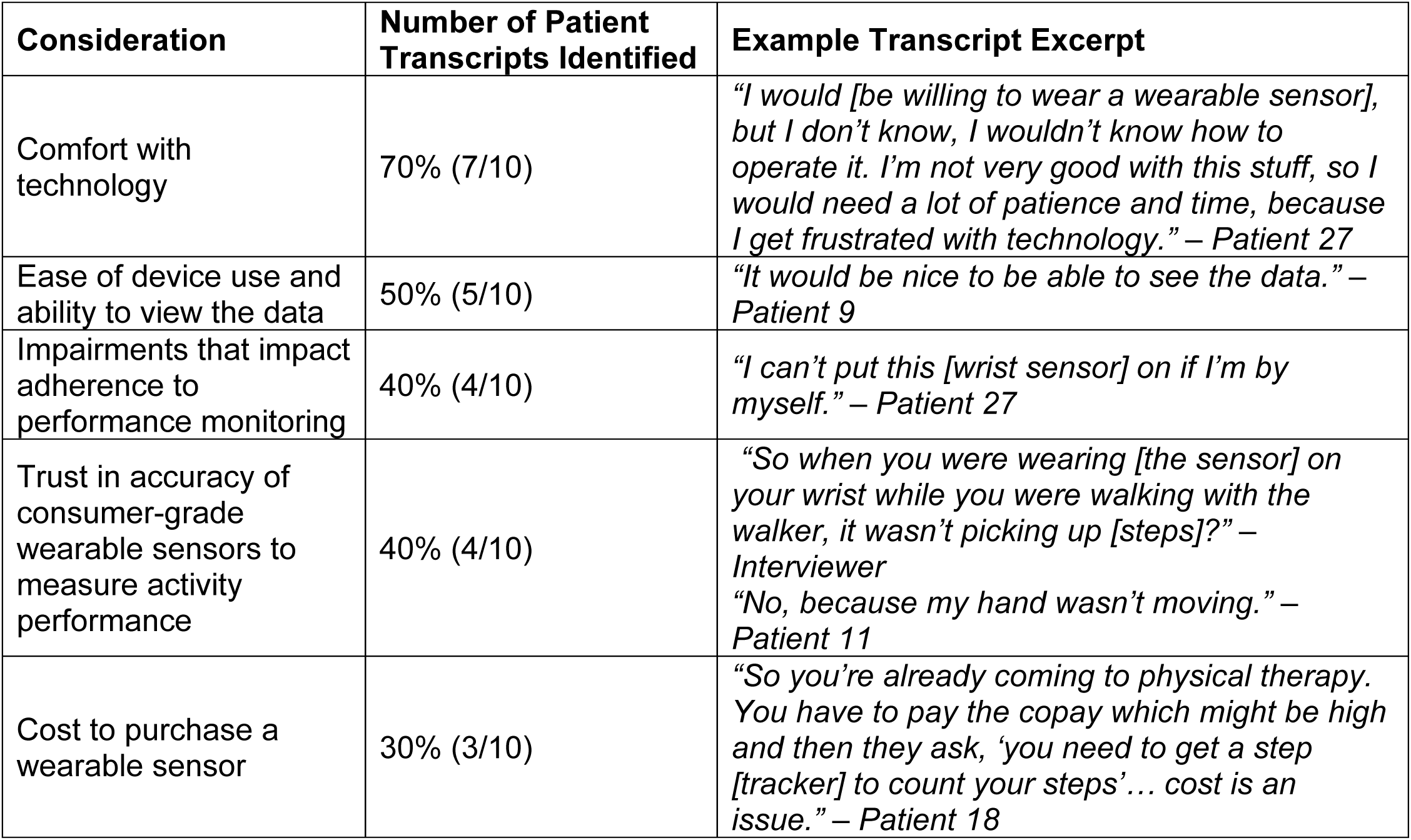
Top Five Patient-Reported Considerations to Integrating Wearable Sensor Technology into Rehabilitation Care.

Despite these considerations, all clinicians interviewed recognized the potential for wearable sensor technology to bridge the gap between activity capacity and activity performance and improve outcomes for individuals seeking rehabilitation care.

> “Measuring capacity and performance is one of the most important things in outpatient therapy, especially because a lot of times we see that the way that a patient is moving or walking or transferring in our clinic can be a lot different than how they’re moving outside of the clinic, especially in the amount that they’re moving outside the clinic… I think that’s why we like to use external devices like step trackers and heart rate monitors…and all those things, because then it’s not necessarily on the patient’s report of how much they’re doing. We actually have concrete evidence and that makes a big difference, and it helps in terms of education, too… I think having ways to assess actual performance makes a big difference and success in outpatient therapy especially.” – Clinician 10

### Research Question 4. What are the data collection and workflow preferences of clinicians and patients for integrating wearable sensor technology into rehabilitation care?

On average, clinicians reported being willing to spend 16.25 ± 7.19 minutes to issue a wearable sensor to a patient, including initializing the sensor to start collecting data, assisting the patient with downloading an associated mobile app, and instructions for wearing and syncing (Figure 2 left panel). The majority (63%) of clinicians reported a preference for continuous monitoring in which they would issue a sensor to their patient within the first few therapy sessions and advise the patient to wear it until discharge from outpatient services (Figure 2, middle panel). All patients reported a willingness to wear a wearable sensor prescribed by their therapist clinician during outpatient rehabilitation care. As some research-based protocols used to measure upper limb activity performance require participants to wear two sensors, one on each wrist,(36, 37) patients were also asked about their willingness to wear sensors on each wrist during outpatient rehabilitation care. All patients reported being willing to do so, with four reporting aesthetic concerns: *“I’m not comfortable wearing devices on both wrists at a social event… Is it okay if I take them off and put them back on when I get home?” – Patient 12.* Data syncing preferences generally fell into two categories: as often as their therapist clinician instructed (60%) or once per day at maximum (40%) (Figure 2, right panel).

**Figure 2.**
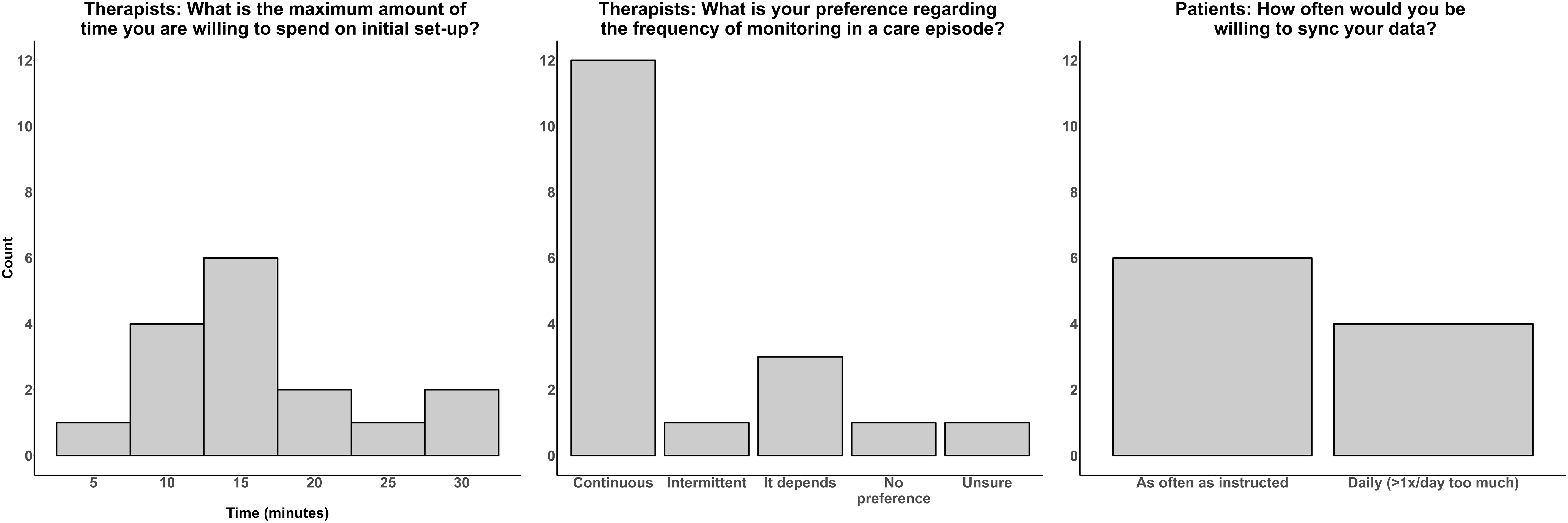
Data Collection Preferences of Clinicians and Patients. The left plot displays a histogram of the maximum number of minutes clinicians were willing to spend on initial set-up of the sensor (mean ± standard deviation: 16.25 ± 7.19 minutes). The middle bar graph displays clinician preferences for the frequency of performance monitoring within a therapy care episode, with continuous monitoring being the most common preference. The right bar graph displays patients’ willingness to sync their data via a mobile app, in which the majority responded they would be willing to do so as often as their therapist clinician instructed.

Interviews with clinicians concluded by discussing possible strategies to best integrate wearable sensor technology into their local clinical workflows. Clinicians identified alternative approaches to integrating wearable sensors into clinical practice which included a clinic inventory of sensors that clinicians could use for activity performance monitoring while the patient was being seen for outpatient rehabilitation services. Several clinicians mentioned that if the clinic were to purchase the same model of sensor, this may minimize the amount of time needed to manage the technology during clinic visits since the sensor would be familiar to clinicians.

> “I’m pretty sure the only way to overcome a lot of these barriers would be to having clinics providing wearable sensors like this, because otherwise, it’s kind of on the patient and they have all these barriers that I’ve talked about that I feel like limit them.” – Clinician 10

Another solution for integrating wearable sensor technology into clinical workflows included the integration of activity performance data into the electronic health record (EHR) for clinicians to easily access.

> “I think that if this [sensor] information could be integrated into the medical record, it would change the game, because all of a sudden it makes it easier for a therapist who has really high expectations of productivity.” – Clinician 6

Challenges with these approaches from the clinician perspective included patients forgetting to sync their data which could result in the data not being available in the EHR, device loss resulting in increased financial burden on the clinic, and concerns about continued monitoring after the patient discharges from therapy services if the device is reclaimed by the clinic. Several suggestions were voiced during patient interviews to help alleviate these issues, including developing mechanisms to generate reminders to sync data, developing detailed user manuals with pictures, creating a help line to contact for technical support if needed, and creating a handout displaying options for consumer grade sensors at various price points for patients to consider purchasing for continued monitoring after discharge.

### Conceptual Model

The thematic analysis conducted in this study led to the development of a conceptual model that describes the workflow and process considerations for activity performance monitoring in outpatient rehabilitation care that takes into consideration themes generated from this analysis (Figure 3). This swim lane diagram highlights the process, pitfalls, and opportunities of the various approaches to activity performance monitoring as described by both patients and clinicians. Each row represents an approach discussed, and each column represents activities occurring in the clinic (dark grey) or in the patient’s free-living environment (light grey). This model provides a visual representation of the data across research questions and provides broader implications for clinical practice and future research.

**Figure 3.**
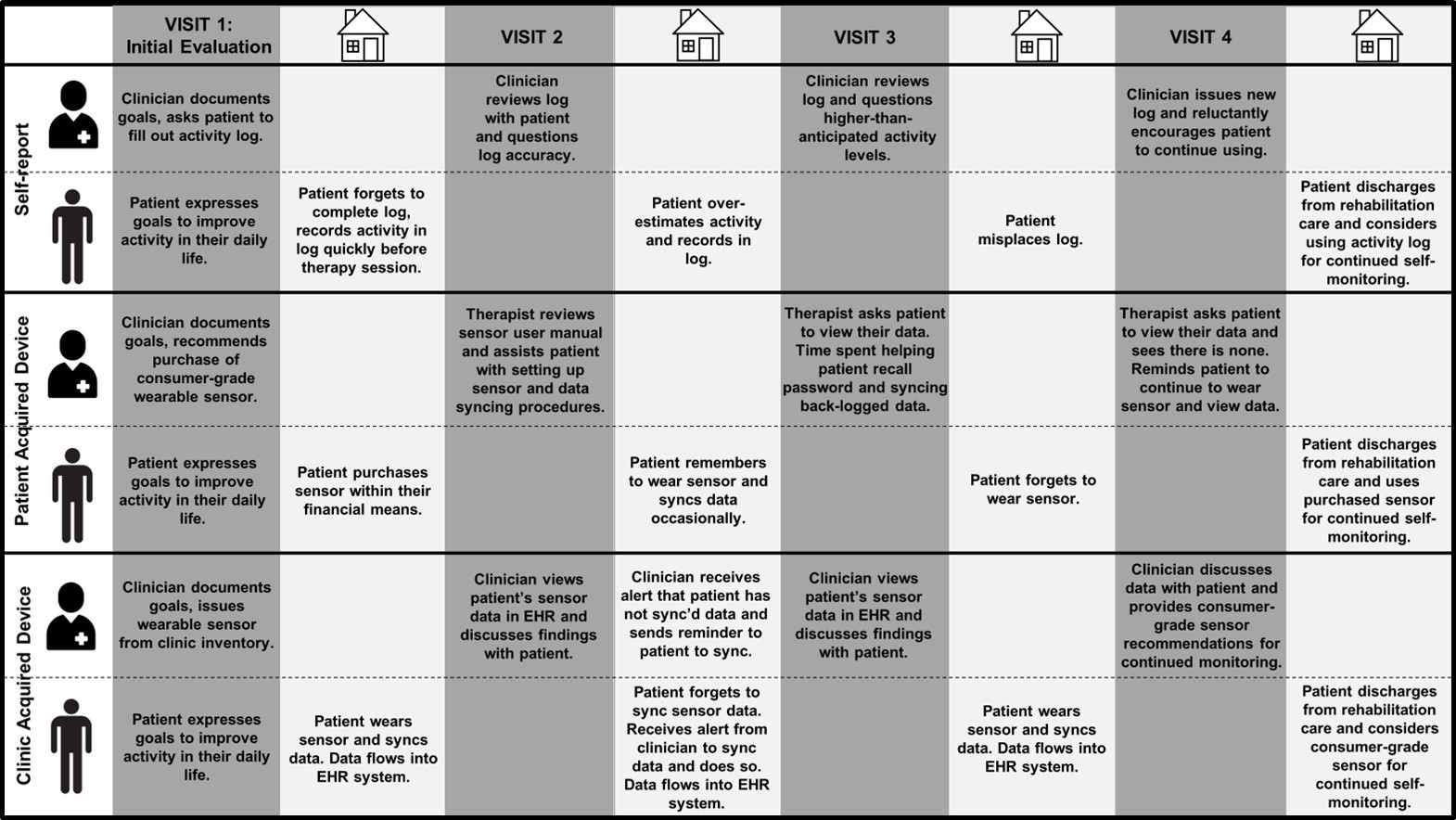
Swim Lanes of Clinicians and Patients During Various Approaches to Activity Performance Monitoring in Rehabilitation. Dark grey columns reflect actions occurring during clinic visits. Lighter grey columns reflect actions occurring outside the clinic while the patient is in their free-living environment. The top third panel displays swim lanes and reported challenges when self-report measures, such as activity logs, are used. The middle third panel displays swim lanes and reported challenges when consumer-grade devices acquired by the patient are used. The bottom third panel displays swim lanes and possible challenges that could occur with a clinic-acquired device model, in which the clinic invests in sensors that clinicians can use for activity performance monitoring in their patients. *Abbreviations: EHR-electronic health record*.

## DISCUSSION

The purpose of this study was to investigate clinician and patient perspectives on integrating wearable sensor technology into rehabilitation care. Using a mixed-methods approach, we found that both clinicians and patients value activity performance monitoring and had preferences for how this monitoring could best be integrated into rehabilitation care. Discussions with clinicians and patients yielded a conceptual model of three different approaches for integrating sensors into rehabilitation care, each with its own barriers and facilitators.

In RQ1, we found that most, but not all, clinicians were able to articulate the distinction between the constructs of activity capacity and activity performance. Improvement in this area could be achieved through reinforcement of these constructs in physical and occupational therapy education programs, or in clinical settings through journal clubs or case presentations that emphasize the constructs of activity capacity and activity performance. Despite this, all clinicians expressed limitations with in-clinic assessments of activity capacity, including sentiments that they do not provide a full picture of the patient and are not necessarily reflective of an individual’s activity performance in daily life. Collectively, these findings suggest that most clinicians are aware of the discrepancy between activity capacity and activity performance in outpatient rehabilitation practice. This was a critical question to investigate since the success of digital health interventions hinges upon a clearly defined problem, which requires stakeholders to both understand and acknowledge the problem.(23) It is important to note, however, that clinicians did not state that in-clinic capacity-based assessments are not important. In-clinic measures of activity capacity are an essential component of rehabilitation practice,(12, 38) predictive of therapy outcomes,(39) and useful for quantifying responses to clinical interventions(40, 41). Thus, the take-away message from RQ1 is not that clinicians felt assessments of activity capacity are not valuable; rather, they perceived activity capacity assessments as a distinct construct from activity performance assessments and that assessments of one construct do not necessarily inform or align with the other in outpatient clinical practice.

In RQ2, we found that most clinicians and patients value activity performance monitoring. As previous studies demonstrate the importance of stakeholder buy-in when implementing new practices (including digital tools) in health care settings, this is a desirable finding.(23, 26) Since not all clinicians and patients felt this was important, however (Figure 1), additional education on the importance of activity performance monitoring will likely be warranted to encourage buy-in. Thus, a pre-implementation strategy for clinics seeking to implement activity performance monitoring could be to measure how much clinicians and patients value performance monitoring and, if needed, provide additional education to facilitate uptake of this practice.(15, 42)

RQ3 generated important considerations for measuring activity performance in outpatient rehabilitation care using two current approaches: self-report and consumer-grade sensors acquired by the patient. Clinicians described barriers with self-report measures that align with those reported in previous work, including recall bias and over- and under-reporting of activity performance compared to sensor-based assessment.(17, 18) Offsetting these barriers is the low cost involved with this approach. Clinics seeking to implement a low-cost approach to activity performance monitoring could consider developing an activity log template and/or having activity calendars readily available for clinicians to issue to their patients to record their activity. This approach can also be helpful if a patient does not have the financial means to purchase a consumer-grade wearable sensor, the alternative approach to activity performance monitoring described by clinicians and patients. Using a consumer-grade sensor acquired by the patient, clinicians and patients appreciated receiving “concrete” data from the sensor but mutually agreed that cost, comfort with using technology, the capabilities of the patient, and accuracy of the sensor need to be considered before pursuing this approach.

Clinics seeking to implement this approach could consider developing a handout of options of consumer grade sensors at various price points for patients to consider, as described by several clinicians. Gathering user manuals of common devices to have on hand for clinicians to refer to may alleviate concerns related to clinician familiarity with the wide variety of consumer sensors available. This wide variety in options has also resulted in varying levels of accuracy across sensors, an important consideration voiced by both clinicians and patients.(43–49) There are various levels of validation to consider when determining whether a sensor is appropriate to use in a specific patient or clinical population, some of which are the responsibility of the manufacturer and some fall on the entity interested in using the sensor (e.g., clinical trial sponsor, researcher, clinician).(50) Clinicians seeking guidance here could consider comparing manual counts to the device output in the clinic to help estimate the accuracy (and overall appropriateness) of a particular device.(51)

In RQ4, clinicians expressed preferences for continuous monitoring and to keep initial set-up time of the sensor to less than ∼16 minutes of therapy session time. Several clinicians justified their preference for continuous monitoring as “more is better” in terms of data generated from the sensor and obtaining a comprehensive picture of the patient’s activity performance in daily life. This approach, however, will need to be weighed against higher participant burden to wear the sensor continuously, the potential of having to purchase a greater number of sensors in the clinic investment model (Figure 3, bottom panel), and larger quantities of data generated that need to be processed and presented in a digestible format to both clinicians and patients. How to best handle the enormous amounts of data generated is an important topic in remote monitoring research, as is understanding the delicate balance between providing clinicians with the appropriate amount of information needed and overwhelming them with streams of unnecessary data.(52) Patient preferences included a willingness to wear one or two sensors, with some aesthetic concerns expressed with the two-sensor approach, and data syncing on a daily basis or as often as instructed by their therapist clinician. Adherence to wearing one or two sensors is generally high in the research setting (53–55); however, compliance with sensor wearing in routine clinical care outside the rigidity of research protocols remains understudied.(56) Thus, future work should consider measuring adherence to wearing (and, if applicable, syncing) sensors in routine rehabilitation care and understanding patient characteristics associated with adherence, as this has been shown to be an important factor in the efficacy of remote monitoring protocols.(22, 57) Elucidating patient characteristics associated with adherence will also inform which patients may need additional supports to participate in remote activity performance monitoring, an important consideration to using wearable sensors voiced by both clinicians and patients.

Discussions with clinicians and patients yielded a conceptual model highlighting three approaches to activity performance monitoring in rehabilitation care, each with barriers and facilitators (Figure 3). The key difference between these approaches can be distilled down to the entity that incurs the cost of a wearable sensor, which may change in the future as wearable sensor technology evolves and the ever-changing landscape of health care reimbursement shifts. Regardless of the approach taken, evidence suggests that new technologies or practices are most likely to be adopted when integrated within clinical workflows.(23, 26, 58) In the context of remote monitoring, integrating data from the technology into the EHR maximizes the probability that the data will be seen and used.(4, 23, 59) Indeed, several clinicians suggested sensor data integration with the EHR to help minimize the time required to access and view their patient’s data. This might be easier to achieve with a consistent device model(s) to minimize challenges related to sensor interoperability and variability in how issues related to patient privacy are managed across sensor vendors. Regardless of how activity performance monitoring is implemented in rehabilitation care, efforts should be made to do so as most patients’ goals are related to improving activity performance in daily life.(1, 2) These findings add to this impetus by demonstrating that both patients and clinicians value activity performance monitoring and view wearable sensor technology as a solution to bridge the wide gap between the goals of patients (improving activity *performance* in daily life) and what is typically measured and intervened on in rehabilitation practice (activity *capacity*).(5)

### Limitations

There are several important limitations to consider when interpreting the results of this work. The clinicians interviewed were employed within an academic medical center and, in many cases, connected to researchers and well-versed in many aspects of rehabilitation research. Conducting this study outside of an academic medical center or at a clinic less familiar with research would have likely generated different findings, such as lower ratings on the perceived importance of activity performance monitoring and less time willing to spend on sensor set-up. Our findings therefore likely represent a more optimistic perspective on integrating wearable sensor technology into rehabilitation care. A related limitation is the small sample size, particularly with the patient cohort. While we reached saturation of information gleaned, the overall number of people queried is small. Thus, the perspectives of clinicians and patients in this study may or may not reflect how clinicians and patients feel about these issues in general. It is unknown how these results generalize to other settings. Efforts to integrate wearable sensor technology (and activity performance monitoring in any form) will likely be setting-specific and ideally built around clinician and patient preferences. Thus, findings from this study may not be informative for efforts to integrate wearable sensor technology at some clinics.

## Conclusions

Rehabilitation clinicians and patients value activity performance monitoring in rehabilitation care and view wearable sensor technology as a solution to bridge the wide gap between the goals of patients (improving activity *performance* in daily life) and what is typically measured and intervened on in rehabilitation practice (activity *capacity*).

These findings serve as a launching point for future studies to investigate the implementation of wearable sensors into rehabilitation care, measure the usability of systems that integrate sensor data into EHR systems, and downstream effects on practice and outcomes. Critically, these initiatives will pave the way for future opportunities to harness the potential of digital technologies to improve the delivery and outcomes of rehabilitation care.

## Conflict of Interest

The authors declare that the research was conducted in the absence of any commercial or financial relationships that could be construed as a potential conflict of interest.

## Author Contributions

AEM: Conceptualization, data curation, formal analysis, funding acquisition, investigation, methodology, project administration, visualization, writing-original draft, writing-review and editing; CLH: conceptualization, data curation, formal analysis, investigation, methodology, supervision, writing-original draft, writing-review and editing; MDB: formal analysis, writing-review and editing; EFC: conceptualization, methodology, writing-review and editing; CAN: methodology, writing-review and editing; TMM: conceptualization, methodology, writing-review and editing; CEL: conceptualization, funding acquisition, methodology, supervision, writing-review and editing.

## Funding

Foundation for Physical Therapy Research Digital Physical Therapy Grant (PI: Miller); Washington University Institute of Clinical and Translational Sciences (ICTS) Just-In-Time Core Usage Funding Program (mHRC-JIT996; PI: Lang); NIH R01/R37 HD068290 (PI: Lang); NIH T32 HD007434 (PI: Lang); NIH K08 MH120341 (PI: Fitzsimmons-Craft)

## Data Availability

The datasets generated and analyzed for this study can be made available upon request.

## Acknowledgements

We are grateful to the clinicians and patients who participated in this study.

## Notes

### Competing Interest Statement

The authors have declared no competing interest.

### Funding Statement

This study was funded by: Foundation for Physical Therapy Research Digital Physical Therapy Grant (PI: Miller); Washington University Institute of Clinical and Translational Sciences (ICTS) Just-In-Time Core Usage Funding Program (mHRC-JIT996; PI: Lang); NIH R01/R37 HD068290 (PI: Lang); NIH T32 HD007434 (PI: Lang); NIH K08 MH120341 (PI: Fitzsimmons-Craft)

### Author Declarations

The Human Research Protection Office of Washington University gave ethical approval for this work.

